# Impact of SARS-CoV-2 variant on the severity of maternal infection and perinatal outcomes: Data from the UK Obstetric Surveillance System national cohort

**DOI:** 10.1101/2021.07.22.21261000

**Authors:** Nicola Vousden, Rema Ramakrishnan, Kathryn Bunch, Edward Morris, Nigel Simpson, Christopher Gale, Patrick O’Brien, Maria Quigley, Peter Brocklehurst, Jennifer J Kurinczuk, Marian Knight

## Abstract

**Background:** In the UK, the Alpha variant of SARS-CoV-2 became dominant in late 2020, rapidly succeeded by the Delta variant in May 2021. The aim of this study was to compare the impact of these variants on severity of maternal infection and perinatal outcomes within the time-periods in which they predominated.

**Methods:** This national, prospective cohort study collated data on hospitalised pregnant women with symptoms of confirmed SARS-CoV-2 infection and compared the severity of infection and perinatal outcomes across the Wildtype (01/03/20-30/11/20), Alpha (01/12/20-15/05/21) and Delta dominant periods (16/05/21-11/07/21), using multivariable logistic regression.

**Findings:** Of 3371 pregnant women, the proportion that experienced moderate to severe infection significantly increased between Wildtype and Alpha periods (24.4% vs. 35.8%; aOR1.75 95%CI 1.48-2.06), and between Alpha and Delta periods (35.8% vs. 45.0%; aOR1.53, 95%CI 1.07-2.17). Compared to the Wildtype period, symptomatic women admitted in the Alpha period were more likely to require respiratory support (27.2% vs. 20.3%, aOR1.39, 95%CI 1.13-1.78), have pneumonia (27.5% vs. 19.1%, aOR1.65, 95%CI 1.38-1.98) and be admitted to intensive care (11.3% vs. 7.7%, aOR1.61, 95%CI 1.24-2.10). Women admitted during the Delta period had further increased risk of pneumonia (36.8% vs. 27.5%, aOR1.64 95%CI 1.14-2.35). No fully vaccinated pregnant women were admitted between 01/02/2021 when vaccination data collection commenced and 11/07/2021. The proportion of women receiving pharmacological therapies for SARS-CoV-2 management was low, even in those critically ill.

**Interpretation:** SARS-CoV-2 infection during Alpha and Delta dominant periods was associated with more severe infection and worse pregnancy outcomes compared to the Wildtype infection, which itself increased risk compared to women without SARS-CoV-2 infection.^1^ Clinicians need to be aware and implement COVID-specific therapies in keeping with national guidance. Urgent action to tackle vaccine misinformation and policy change to prioritise uptake in pregnancy is essential.

**Funding:** National Institute for Health Research HS&DR Programme (11/46/12).

## INTRODUCTION

In 2020 the World Health Organisation’s (WHO) living systematic review concluded that SARS-CoV-2 infection during pregnancy was associated with an increased risk of admission to intensive care (ICU) for the mother, increased risk of preterm birth and admission for neonatal care for the infant.^2^ Included studies predominantly contained data from the USA and China and were conducted in the first six months of the pandemic, prior to the spread of new variants.

In the UK, a new variant of SARS-CoV-2 (B.1.1.7, Alpha Variant of Concern (VOC)) was initially reported in South East England in September 2020 and then circulated at very low levels in the population until mid-November 2020 when it then dominated.^3^ This was then succeeded by the Delta VOC (B.1.617.2) which quickly became the dominant variant in late May 2021.^4^ There is growing evidence that in the non-pregnant population, the Alpha VOC may be associated with increased risk of hospitalisation and mortality compared with other lineages.^5^ Most recently, data from a Scottish national cohort demonstrated that infection with the Delta VOC approximately doubled the risk of hospital admission in the general population, compared to infection with the Alpha VOC.^6^ However, there are very limited published studies exploring the impact of different SARS-CoV-2 variants on pregnancy and perinatal outcomes.

A single centre study from the UK reported a significant increase in peripartum referrals for extracorporeal membrane oxygenation (ECMO) during the second wave of the pandemic, when the Alpha VOC became dominant (n=19 vs n=4)^7^. This was in keeping with findings of a national registry of patients admitted to ICU, which reported an increase in the number of pregnant or recently pregnant women in the second wave compared to the first.^8^ However, these reports were limited by the absence of a comparator, meaning it was not possible to determine whether this was a result of changing variants as opposed to an increasing total numbers of infected women.

To the best of our knowledge, only two further publications have explored the potential impact of different SARS-CoV-2 variants in pregnancy. A retrospective cohort from a single centre in India concluded that pregnant women admitted during the Delta VOC dominant second wave (n=387) had higher rates of admission to ICU or high dependency unit (11.6 vs 2.4%) and case fatality (5.7 vs. 0.7%) than those in the first wave (n=1143).^9^ This is in keeping with a review of 803 maternal deaths with SARS-CoV-2 in Brazil, where a significantly higher case fatality rate was reported in 2021 (Gamma VOC) compared to 2020 (15.6% vs. 7.4%).^10^ These preliminary studies suggest an urgent need for robust national data on the impact of new variants on maternal and perinatal outcomes in order to inform policy.

The primary aim of this study was therefore to compare the impact of SARS-CoV-2 infection on severity of maternal infection and perinatal outcomes across three time periods in which the Wildtype, Alpha and Delta VOCs were dominant.

## METHODS

### Data sources

A national, prospective observational cohort study was conducted using the UK Obstetric Surveillance System (UKOSS).^11^ UKOSS is a research platform that was established in 2005. All 194 hospitals in the UK with a consultant-led maternity unit collect population-based information about specific severe pregnancy complications. Nominated reporting clinicians, facilitated by research midwives and nurses from the UK’s National Institute of Health Research Clinical Research Network, notified all pregnant women admitted to their hospital with confirmed SARS-CoV-2 infection. In addition to receipt of real-time reports, zero reports were confirmed. Reporters who had notified a case but not returned data received email reminders. Hospital admission was defined as an overnight hospital stay, or longer, for any cause, or admission of any duration to give birth. Women were taken as confirmed SARS-CoV-2 if they were hospitalised during pregnancy or within two days after giving birth and had a positive test during or within seven days of admission. Women not meeting this case definition, and those without any symptoms of SARS-CoV-2 infection, were excluded (Supplementary Figure 1). Information on women who died, or who had stillbirths or neonatal deaths, was cross-checked with data from the organisation responsible for maternal and perinatal death surveillance in the UK (MBRRACE-UK).^12^

### Measures

The primary outcome was a composite indicating moderate to severe SARS-CoV-2 infection: oxygen saturation <95% on admission, need for oxygen therapy, evidence of pneumonia on imaging, admission to ICU or maternal death, based on the WHO criteria of COVID-19 disease severity^13^. Each of those components was also analysed separately, as were pregnancy and perinatal outcomes including mode and gestation of birth, stillbirth, live birth, admission to neonatal intensive care and neonatal death.

As individual-level SARS-CoV-2 variant data were not recorded in medical records, the outcomes were compared across three proxy groups according to the time-period in which three different SARS-CoV-2 variants were the dominant circulating strain in the UK. The original ‘Wildtype’ period included women admitted to hospital from 1^st^ March to 30^th^ November 2020, the Alpha period, from 1^st^ December 2020 to 15^th^ May 2021, and the Delta period, from 16^th^ May 2021 to 11^th^ July 2021. Cut-offs for the Delta period were chosen using data on variant sequencing from Public Health England to identify the week that this variant first contributed more than 50% of cases nationally.^4^

### Study registration

The study was registered with ISRCTN, number 40092247 and the protocol is available at https://www.npeu.ox.ac.uk/ukoss/current-surveillance/covid-19-in-pregnancy.

### Ethics and consent

This study was approved by the HRA NRES Committee East Midlands – Nottingham 1 (Ref. Number: 12/EM/0365).

### Statistical methods and analysis

Statistical analyses were performed using STATA version 15 (Statacorp, TX, USA). Numbers and proportions are presented with 95% confidence intervals (CI). Where data were missing, proportions are presented out of cases known. Odds ratios (ORs) with 95% CI were estimated using unconditional logistic regression.

The hypothesised relationships between SARS-CoV-2 variant and severity of infection were identified using directed acyclic graphs, created with DAGitty.net.^14,15^ (Supplementary Figure 2). These were informed by associations identified in the literature and underlying theory. The minimum adjustment set to control for confounding bias was sociodemographics (age, ethnicity, body mass index (BMI) and employment), and vaccine status. However, there were insufficient data to include vaccine status as a covariate (as data were only collected from 01/02/2021) and therefore, based on the DAG, it was necessary to also include pre-existing medical conditions (asthma, cardiac disease, diabetes or hypertension) in order to block a further potential biasing pathway (Supplementary Figure 2).^1^ These were included in the model as a combined covariate if any of the conditions were identified. In the absence of data sparsity or multicollinearity (highest Spearman correlation coefficient of 0.19), all pre-specified covariates as identified by the DAG were included. Following testing for departure from linearity using likelihood ratio testing, age and BMI were included as ordered categorical variables. Potential effect modifiers were identified *a priori* as the covariates identified in the DAG, in addition to parity and trimester of pregnancy at time of infection. Plausible interactions were tested by the addition of interaction terms and subsequent likelihood ratio testing on removal, with a p-value <0.01 considered as evidence of significant interaction. No interaction terms were included in the model. In this national observational study, the study sample size was governed by the disease incidence, thus no formal power calculation was carried out.

## RESULTS

In total, 3371 women were admitted to hospital across the UK with symptoms of confirmed SARS-CoV-2 infection between 1^st^ March 2020 and 11^th^ July 2021. Most cases were during the second wave when the Alpha VOC was dominant (Figure 1). Of those where the primary reason for admission was known (74.8%, n=2521), just under half (45.0%, n=1137) were admitted for COVID-19, 30.0% (n=755) for labour and birth and 25.0% (n=629) for other obstetric reasons. The proportion admitted primarily for COVID-19 increased across the variants from 41.4% (n=204), to 45.9% (n=384) and 54.2% (n=90) in the Wildtype, Alpha and Delta periods, respectively. The characteristics of each group are described in Table 1. The proportion of women admitted during the Delta period aged 35 years or over was 22.9% (n=39) compared to 29.5% (n=422) and 28.8% (n=508) in Wildtype and Alpha periods respectively. Two thirds reported that they or their partner were in paid employment in the Delta period (62.6% (n=107)) compared to nearly 80% in the Alpha and Wildtype periods (79.6% (n=1142) and 75.8% (n=1338) respectively). Across all three periods, the majority of women were overweight or obese. The proportion of women admitted in the Alpha and Delta periods with one or more pre-existing medical conditions was 14.0%, (n=247) and 13.5% (n=23), compared to 11.8% (n=169) of those admitted in the Wildtype period. The most common time for admission was at term across all three time periods.

**Figure 1:**
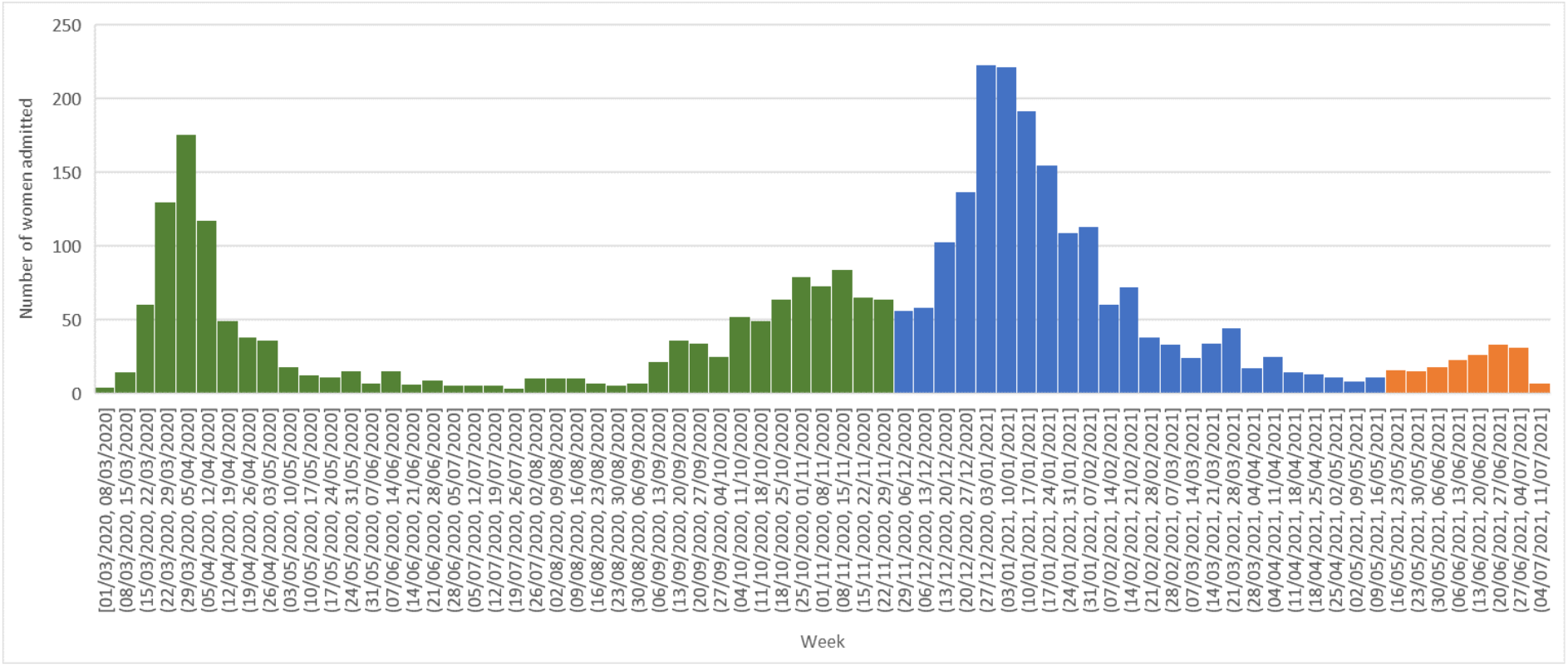
Admissions of pregnant women with symptomatic confirmed SARS-CoV-2 to UK hospitals during Wildtype (01/03/20-30/11/2 2020, Green), Alpha (01/12/20 – 15/05/21, Blue) and Delta periods (16/05/21-11/07/21, Orange)

**Table 1:** Characteristics of pregnant women with confirmed symptomatic SARS-CoV-2 infection admitted to hospital in the UK during the periods in which the Wildtype, Alpha and Delta variants were dominant.

| Characteristic | Wildtype | Alpha | Delta |
| --- | --- | --- | --- |
| **Age (years):** | **N=1435 (%)** | **N=1765 (%)** | **N=171 (%)** |
| <20 | 20 (1.4) | 18 (1.0) | 4 (2.4) |
| 20-34 | 991 (69.2) | 1236 (70.2) | 127 (74.7) |
| ≥35 | 422 (29.5) | 508 (28.8) | 39 (22.9) |
| Missing | 2 | 3 | 1 |
| **Body Mass Index (BMI) (kg/m²):** |  |  |  |
| Underweight (<18.5) | 21 (1.5) | 22 (1.3) | 0 (0) |
| Normal (18.5 to <25) | 456 (33.1) | 564 (33.6) | 59 (37.1) |
| Overweight (25 to <30) | 444 (32.2) | 486 (28.9) | 48 (30.2) |
| Obese (>30) | 458 (33.2) | 609 (36.2) | 52 (32.7) |
| Missing | 56 | 84 | 12 |
| **Either woman or partner in paid work** | 1142 (79.6) | 1338 (75.8) | 107 (62.6) |
| **Ethnic Group** |  |  |  |
| White | 707 (50.4) | 1012 (58.8) | 83 (51.2) |
| Asian | 418 (29.8) | 411 (23.9) | 51 (31.5) |
| Black | 177 (12.6) | 179 (10.4) | 16 (9.9) |
| Chinese/Other | 71 (5.1) | 71 (4.1) | 9 (5.6) |
| Mixed | 31 (2.2) | 37 (2.7) | 3 (1.9) |
| Missing | 31 | 45 | 9 |
| **Current smoking** | 98 (7.2) | 147 (8.6) | 14 (8.8) |
| Missing | 67 | 46 | 12 |
| **Pre-existing medical conditions** |  |  |  |
| Asthma | 91 (6.3) | 178 (10.1) | 15 (8.8) |
| Hypertension | 40 (2.8) | 35 (2.0) | 5 (2.9) |
| Cardiac disease | 20 (1.4) | 25 (1.4) | 2 (1.2) |
| Diabetes | 39 (2.7) | 30 (1.7) | 2 (1.2) |
| **Gestational Diabetes** | 147 (10.2) | 184 (10.4) | 19 (11.1) |
| **Multiparous** | 857 (60.2) | 1147 (65.5) | 105 (64.8) |
| Missing | 11 | 15 | 9 |
| **Multiple pregnancy** | 36 (2.5) | 35 (2.0) | 1 (0.6) |
| **Gestation at admission (weeks)** |  |  |  |
| <22 | 142 (10.0) | 157 (9.0) | 18 (10.7) |
| 22-27 | 159 (11.2) | 207 (11.9) | 31 (18.3) |
| 28-31 | 190 (13.3) | 213 (12.2) | 24 (14.2) |
| 32-36 | 340 (23.9) | 452 (25.9) | 42 (24.9) |
| 37 or more | 594 (41.7) | 717 (41.1) | 54 (32.0) |
| Missing | 10 | 19 | 2 |

Out of 742 women where vaccine status was collected (n=571 during the Alpha period and n=171 during the Delta period), a total of four women admitted with symptomatic SARS-CoV-2 infection had received their first dose of SARS-CoV-2 vaccination prior to their positive test (from 5 to 16 weeks prior). One was admitted during the Alpha period (0.2% of women) and three during the Delta period (1.8% of women). One woman was recorded as vaccinated but was missing a date and therefore it was not possible to confirm that this preceded infection. There were no women admitted with symptoms of SARS-CoV-2 in this study period that had received both doses of vaccine.

Overall, 25% of women admitted with symptomatic COVID-19 during the Wildtype period had at least one marker of moderate to severe infection. This significantly increased to 35.8% during the Alpha period (aOR 1.75, 95% CI 1.48-2.06) and was greater still during the Delta period when nearly half of women had moderate to severe infection (45.0%, aOR 1.53, 95% CI 1.07-2.17, for Alpha vs Delta periods)(Table 2). There was a total of 15 maternal deaths in women with COVID-19, 10 during the Wildtype and five during the Alpha period (Table 2). After adjustment, women admitted during the Alpha period were significantly more likely to require admission to ICU than those admitted during the Wildtype period (11.3% vs. 7.7%, aOR 1.61, 95% CI 1.24-2.10). There was also a statistically non-significantly increased risk of ICU admission in women being admitted during the Delta compared to Alpha periods (15.2% vs. 11.3%, aOR 1.60, 95% CI 0.99-2.59).

**Table 2:** Respiratory and medical support of pregnant women symptomatic of SARS-CoV-2 during the periods in which the Wildtype, Alpha and Delta variants were dominant.

|  | Wildtype<br>N=1435<br>(%) | Alpha<br>N=1765<br>(%) | Delta<br>N=171<br>(%) | OR Alpha<br>vs.<br>Wildtype<br>(95% CI) | aOR Alpha<br>vs.<br>Wildtype<br>(95% CI) | OR<br>Delta<br>vs.<br>Alpha<br>(95%<br>CI) | aOR<br>Delta<br>vs.<br>Alpha<br>(95%<br>CI) |
| --- | --- | --- | --- | --- | --- | --- | --- |
| **Composite indicator of moderate to severe infection** | 350<br>(24.4) | 631<br>(35.8) | 77<br>(45.0) | 1.72 (1.48-2.01) | 1.75 (1.48-2.06) | 1.47 (1.07-2.02) | 1.53 (1.07-2.17) |
| **Oxygen saturation measured on admission (Yes)** | 539<br>(37.6) | 1255<br>(71.1) | 132<br>(77.2) | NC | NC | NC | NC |
| Median Oxygen saturation (IQR) | 98 (96-99) | 97 (96-98) | 97 (96-99) | NC | NC | NC | NC |
| Oxygen saturation <95% | 54 (3.8) | 185 (10.5) | 17 (9.9) | NC | NC | NC | NC |
| **Evidence of pneumonia on imaging** | 274<br>(19.1) | 486<br>(27.5) | 63<br>(36.8) | 1.61(1.36-1.90) | 1.65 (1.38-1.98) | 1.54 (1.12-2.13) | 1.64 (1.14-2.35) |
| **Respiratory support required** | 183<br>(20.3) | 466<br>(27.2) | 52<br>(33.3) | 1.47 (1.21-1.78) | 1.39 (1.13-1.71) | 1.34 (0.95-1.90) | 1.43 (0.97-2.11) |
| Non-invasive oxygen (nasal canulae, mask or non-rebreathe mask at <15l/min) | 107<br>(61.1) | 292<br>(64.8) | 29<br>(69.1) | REF | REF | REF | REF |
| High flow oxygen (>15l/min) or CPAP | 28 (16.0) | 71 (15.7) | 11 (26.2) | 0.93 (0.57-1.52) | 1.04 (0.61-1.77) | 1.56 (0.74-3.27) | 2.01 (0.91-4.44) |
| Invasive Ventilation or ECMO | 40 (22.9) | 88 (19.5) | 2 (4.5) | 0.81 (0.52-1.24) | 0.99 (0.61-1.59) | 0.23 (0.05-0.98) | 0.30 (0.07-1.30) |
| Level not known | 8 | 15 | 10 | - | - | - | - |
| **Critical Care received** | 111 (7.7) | 199 (11.3) | 26 (15.2) | 1.52 (1.19-1.94) | 1.61 (1.24-2.10) | 1.41 (0.91 - 2.20) | 1.60 (0.99-2.59) |
| **Maternal Death** | 10 (0.7) | 5 (0.3) | 0 (0) | NC | NC | NC | NC |
| **Pharmacological Management Total\*** | 99 (6.9) | 253 (14.3) | 28 (16.3) | 2.26 (1.77-2.88) | 2.37 (1.83-3.07) | 1.17 (0.76-1.79) | 1.35 (0.87-2.12) |
| **Antivirals Total** | 43 (3.0) | 33 (1.9) | 3 (1.8) | NC | NC | NC | NC |
| **Tocilizumab** | 0 (0) | 22 (1.3) | 7 (4.1) | NC | NC | NC | NC |
| **Steroids for maternal indication** | 68 (4.7) | 219 (12.4) | 25 (14.6) | NC | NC | NC | NC |
| **Regeneron Monoclonal Antibodies** | 0 (0) | 6 (0.3) | 0 (0) | NC | NC | NC | NC |
| **Recruited to RECOVERY** | 21 (1.5) | 87 (4.9) | 0 (0) | NC | NC | NC | NC |
| **Steroids for fetal lung maturation** | 252 (17.6) | 336 (19.0) | 26 (15.2) | NC | NC | NC | NC |

\* Any of the listed medications given for medical management of SARS-CoV-2: Antivirals, Tocilizumab, maternal steroids, monoclonal antibodies. NC=not compared. IQR = interquartile range. CPAP = continuous positive airway pressure. ECMO = extracorporeal membrane oxygenation.

Women admitted during the Alpha period were more likely to have SARS-CoV-2 pneumonia confirmed on imaging (27.5% vs 19.1%; aOR 1.65, 95% CI 1.38-1.98) and require respiratory support (27.2% vs. 20.3%; aOR 1.39 95% CI 1.13-1.71) than those admitted in the Wildtype period (Table 2). Furthermore, women admitted during the Delta period were at greater risk again of pneumonia compared to the Alpha period, with more than a third having SARS-CoV-2 pneumonia (36.8% vs. 27.5%; aOR 1.64, 95% CI 1.14-2.35) and a third of women requiring respiratory support (33.3% vs. 27.2%; aOR 1.43, 95% CI 0.97-2.11). Whilst not statistically significant, there also appeared to be reduced use of invasive ventilation and increased use of high flow oxygen and continuous positive airway pressure (CPAP) over time.

The proportion that received any pharmacological treatment for COVID-19 (one or more of an antiviral, Tocilizumab, maternal steroids and monoclonal antibodies) was small, but did increase over time: 6.9% in Wildtype vs. 14.3% in Alpha period (aOR 2.37; 95% CI 1.83-3.07) and 16.3% in Delta period (aOR 1.35, 95% CI 0.87-2.12). A greater proportion of women admitted to ICU received any pharmacological therapy for COVID-19 than those not admitted to ICU (39.9%, n=134 vs. 8.1% n=246), although this proportion was still small: 12.5% (n=42) received antivirals, 8.0% (n=27) received Tocilizumab, 27.1% (n=91) received maternal steroids, and 0.6% (n=2) received monoclonal antibodies.

Of those with complete outcome information (96.9% Wildtype, 89.1% Alpha and 42.7% Delta), the median gestation at birth was the same across periods (Table 3). In the Alpha period, 1.4% (n=22) gave birth at between 22 and <28 weeks’ compared to 0.7% (n=10) in the Wildtype period (aOR 2.15, 95% CI 0.97-4.76). The proportion that gave birth at 28 to <32 weeks’ was similar between these periods (3.7% (n=57) vs. 3.3% (n=45), aOR 1.20, 95% CI 0.79-1.83). Given that 10.1% of women in the Alpha period had incomplete delivery information compared to 3.1% in the Wildtype period, a sensitivity analysis assuming the remainder delivered at term was undertaken and demonstrated similar results (22 to <28 weeks aOR 1.99, 95% CI 0.94-4.23; 28 to <32 weeks aOR1.15, 95% CI 0.77-1.71). Since, as anticipated due to timing of this analysis, fewer pregnancies were completed in the Delta period, formal comparison of the proportion of preterm births was not performed.

**Table 3:** Pregnancy outcomes for women symptomatic of SARS-CoV-2 during the periods in which the Wildtype, Alpha and Delta variants were dominant.

| Pregnancy outcomes | Wildtype | Alpha | Delta | OR Alpha vs. Wildtype (95% CI) | aOR Alpha vs. Wildtype (95% CI) | OR Delta vs. Alpha (95% CI) | aOR Delta vs. Alpha (95% CI) |
| --- | --- | --- | --- | --- | --- | --- | --- |
|  | **N=1435 (%)** | **N=1765 (%)** | **N=171 (%)** |  |  |  |  |
| **Ongoing pregnancy** | 44 (3.1) | 193 (10.9) | 98 (57.3) | NC | NC | NC | NC |
| **Pregnancy Loss** | 33 (2.4) | 29 (1.8) | 1 (1.4) | NC | NC | NC | NC |
| **Birth** | 1358 (94.5) | 1543 (87.4) | 72 (42.1) | NC | NC | NC | NC |
| **Gestation at birth (weeks)\*** |  |  |  |  |  |  |  |
| 22-27 | 10 (0.7) | 22 (1.4) | 0 (0) | 1.99 (0.94-4.23) | 2.15 (0.97-4.76) | NC | NC |
| 28-31 | 45 (3.3) | 57 (3.7) | 6 (8.3) | 1.15 (0.77-1.71) | 1.20 (0.79-1.83) | NC | NC |
| 32-36 | 193 (14.2) | 242 (15.7) | 12 (16.7) | 1.14 (0.93-1.40) | 1.15 (0.93-1.43) | NC | NC |
| 37 or more | 1092 (80.4) | 1206 (78.2) | 53 (73.6) | REF | REF | REF | REF |
| Median (IQR) | 39 (37-40) | 39 (37-40) | 39 (37-40) | 0.97 (0.95-1.00) | 0.97 (0.94-1.00) | 0.99 (0.91-1.07) | 0.97 (0.90-1.05) |
| Missing | 18 | 16 | 1 | - | - | - | - |
| **Delivery expedited due to COVID-19** | 87 (11.7) | 204 (13.9) | 17 (25.4) | 1.22 (0.93-1.59) | 1.15 (0.87-1.53) | NC | NC |
| Missing | 617 | 78 | 5 | - | - | - | - |
| **Mode of birth** |  |  |  |  |  |  |  |
| Pre-labour Caesarean | 431 (32.0) | 561 (36.7) | 30 (42.3) | NC | NC | NC | NC |
| Caesarean after labour onset | 201 (14.9) | 211 (13.8) | 6 (8.5) | NC | NC | NC | NC |
| Operative vaginal | 150 (11.2) | 139 (9.1) | 8 (11.3) | NC | NC | NC | NC |
| Unassisted vaginal | 563 (41.9) | 616 (40.3) | 27 (38.0) | NC | NC | NC | NC |
| Missing | 13 | 16 | 1 | - | - | - | - |

\* Excluding pregnancy loss. NC=not compared. IQR = interquartile range.

**Table 4:**
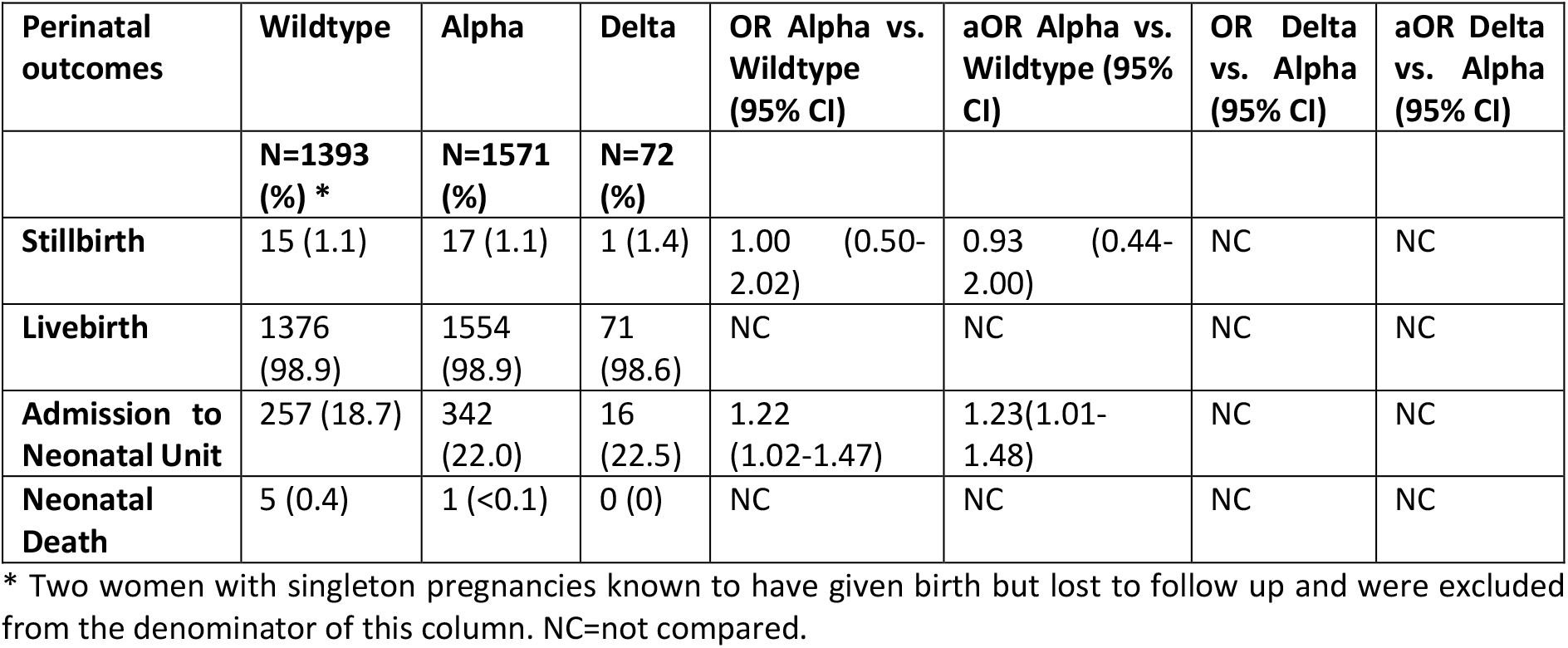
Perinatal outcomes for women symptomatic of SARS-CoV-2 during the periods in which the Wildtype, Alpha and Delta variants were dominant.

| Perinatal outcomes | Wildtype | Alpha | Delta | OR Alpha vs. Wildtype (95% CI) | aOR Alpha vs. Wildtype (95% CI) | OR Delta vs. Alpha (95% CI) | aOR Delta vs. Alpha (95% CI) |
| --- | --- | --- | --- | --- | --- | --- | --- |
|  | **N=1393 (%) \*** | **N=1571 (%)** | **N=72 (%)** |  |  |  |  |
| **Stillbirth** | 15 (1.1) | 17 (1.1) | 1 (1.4) | 1.00 (0.50-2.02) | 0.93 (0.44-2.00) | NC | NC |
| **Livebirth** | 1376 (98.9) | 1554 (98.9) | 71 (98.6) | NC | NC | NC | NC |
| **Admission to Neonatal Unit** | 257 (18.7) | 342 (22.0) | 16 (22.5) | 1.22 (1.02-1.47) | 1.23(1.01-1.48) | NC | NC |
| **Neonatal Death** | 5 (0.4) | 1 (<0.1) | 0 (0) | NC | NC | NC | NC |

\* Two women with singleton pregnancies known to have given birth but lost to follow up and were excluded from the denominator of this column. NC=not compared.

The majority of babies were live born with no change in the proportion of stillbirths across the time periods. There were six neonatal deaths, five in the Wildtype period and one in the Alpha period, none of which were directly related to neonatal SARS-CoV-2 infection. Overall, nearly one in five babies were admitted for neonatal care, with significantly increased risk in those born to mothers admitted in the Alpha compared to Wildtype period (22.0% vs. 18.7%, aOR1.23, 95% CI 1.01-1.48).

## DISCUSSION

This national prospective cohort study has identified that, after adjusting for sociodemographics and pre-existing medical conditions, the proportion of symptomatic pregnant women admitted who experienced moderate to severe COVID-19 has significantly increased from 24% to 36% and then 45% in the Wildtype, Alpha, and Delta periods respectively. Women admitted in the Alpha period were more likely to require respiratory support, have pneumonia, and be admitted to ICU, compared to women admitted in the Wildtype period. Women admitted during the Delta period had a further increase in risk compared to those admitted in the Alpha period, with a greater proportion having pneumonia and non-significant increases in respiratory support and ICU admission. Whilst the majority of babies were live born, babies born to mothers in the Alpha period were more likely to require admission for neonatal care compared to during the Wildtype period.

To our knowledge, this is the first national prospective cohort to compare pregnancy and perinatal outcomes by time-period according to different dominant SARS-CoV-2 variants. A key strength of these data is the existing mechanism for national case identification of all women admitted to hospital. In the UK universal SARS-CoV-2 testing for all obstetric admissions was implemented from May 2020. It is therefore a further strength that this study was restricted to those with symptomatic infection in order to minimise bias associated with universal screening. Women presenting to hospital are inherently more likely to have an adverse outcome and therefore increased adverse outcomes may be incorrectly attributed to SARS-CoV-2 rather than misclassification bias, which impacts most non-population based studies.^16^ Whilst it is a limitation that women with mild infection diagnosed and treated in the community will not be included in this study, it is highly likely that all women with severe infection would have been captured.

A further limitation of our study is that variant sequencing data were not available for individual women, therefore proxy time periods were utilised instead. However, the Delta VOC is now known to have contributed more than 90% of all sequenced cases since 7^th^ June 2021 so major contamination is unlikely.^17^ Other time-dependent changes will exist which we cannot account for, for example varying thresholds for admission to hospital or ICU depending on clinician familiarity with managing COVID-19. In the general population, national guidance was updated in January 2021 to inform community management of those with oxygen saturations >92%.^18^ However, it is unclear that this admission threshold was used extensively in pregnancy and given that the RCOG has never released national admission guidance for pregnant patients, this is unlikely to account for differences observed. Differing thresholds based on bed capacity may have been a contributory factor during the peak of the Alpha VOC when hospital pressures may have restricted admission to the most severe cases. However, this is not supported by our finding of an increased proportion of admissions primarily for COVID-19 in this time compared to the Wildtype period. In addition, current hospital pressures from COVID-19 (Delta VOC dominant) are not reported to be as high as during the second wave,^19^ therefore this could explain the greater proportion of women admitted for COVID-19 in this period, but it does not explain the increase in severe outcomes observed in this study.

We have reported a potential change in the proportion of pregnant women in paid employment between periods. This may be a result of increased unemployment during this period, or an increased proportion of women from more deprived socioeconomic backgrounds. It is a limitation of this study that further information on socioeconomic circumstances could not be collected due to ethics committee requirements. This is important when considering whether disease severity can be attributed to the variant because the variant also impacts on disease transmission. For example, Alpha VOC has been shown to have a higher secondary attack rate and therefore factors that increase transmission, such as multi-occupancy housing and public-facing occupations, are important.^5^ Since socioeconomic deprivation is also a known independent risk factor for adverse pregnancy outcome, this could be a source of residual confounding in this study.

COVID-19-specific pharmacological therapies, which are now standard care, were used infrequently, even for women that were critically unwell. Based in the interim report from the RECOVERY trial, The Royal College of Obstetricians and Gynaecologists (RCOG) recommended in June 2020 that corticosteroid therapy should be considered for all women who were clinically deteriorating.^20^ Whilst usage of steroids has improved, it remains low at 14.6% during the Delta period, and whilst it was double in those critically unwell (30.8% during the Delta period) this still represents a small proportion of pregnant women being treated appropriately. The recent confidential enquiry (MBRRACE-UK)^20^ into care of all pregnant and postnatal women who died with SARS-CoV-2 found that only one in ten had received treatment in accordance with the evidence-based guidance. This study highlights that pregnant woman are at increasing risk of severe disease from SARS-CoV-2.^21^ Health care professionals need to be alert to the risk of deterioration and initiate management for all women in line with national guidance. Pregnant women need to be reassured about the availability and safety of effective treatments and advised to avoid delay in seeking care.

Vaccination for all pregnant women regardless of risk group in the UK was recommended by the Joint Committee on Vaccination and Immunisation (JCVI) on 16^th^ April 2021.^22^ Prior to this, vaccination has been available to pregnant women with underlying health conditions or increased risk of exposure since 31^st^ December 2020.^23^ National data from Scotland suggests that vaccine update in pregnancy is very low, with 2% of the 3603 women that delivered in May 2021 having any vaccine dose.^24^ Public Health England have also recently reported that to date, 51,724 pregnant women in England have received their first dose, and of these 20,648 are fully immunised, where approximately 643,000 women give birth each year.^22^ Whilst it is greatly reassuring that in our study there were no fully vaccinated pregnant women admitted with symptomatic SARS-CoV-2, it is a limitation that there were insufficient data to examine the impact of vaccination status on severity of infection. A survey undertaken by the RCOG in May 2021 reported that of 844 pregnant women offered vaccination, 58% had declined, predominantly due to fear over safety for the mother and baby.^25^ There has been widespread misinformation regarding the safety of the vaccination in young women^26^, likely fuelled by changing advice on the safety of vaccines in pregnant women when they first became available. The findings of this study strongly highlight the urgent need for an international approach to tackle this misinformation and improve uptake of the vaccine during pregnancy, potentially through change of policy to prioritise appointments for pregnant women and bring forward second doses. This is of even greater importance as Delta VOC continues to rapidly rise in both high and low-resourced settings.^27^

In conclusion, this national study has demonstrated that pregnant women admitted during the periods in which the Alpha VOC and Delta VOC are dominant, are at increased risk of moderate to severe COVID-19, resulting in admission to ICU. This is against the background of an already increased risk compared to the pregnant population without SARS-CoV-2.^1^ Effective treatments are now available, but are used in only a minority of cases, even amongst those that are critically unwell. Healthcare professionals need to be aware of the increased risk of deterioration observed with Delta VOC and increase utilisation in keeping with national guidance. The absence of admission in pregnant women that have been fully vaccinated against SARS-CoV-2 supports the effectiveness of immunisation, yet vaccine uptake is reported to be low compared to the general population. Urgent action to tackle misinformation and policy change to prioritise actions to promote uptake are required, given the increasing rates of Delta VOC nationally and internationally.^28^

## Data Availability

Data cannot be shared publicly because of confidentiality issues and potential identifiability of sensitive data as identified within the Research Ethics Committee application/approval. Requests to access the data can be made by contacting the National Perinatal Epidemiology Unit data access committee via.

## Author Contributions

All authors contributed to conceptualisation, the writing and editing of this study, had final approval of the version to be published and agree to be accountable for all aspects of the work. KB, EM, NS, CG, PO, MQ, PB, JK and MK contributed to funding acquisition, supervision, and methodology. NV, RR, KB and MK contributed to data curation and formal analysis, and all had access to verify the underlying data.

## Acknowledgements

The study was funded by the National Institute for Health Research HS&DR Programme (project number 11/46/12). MK is an NIHR Senior Investigator. The views expressed are those of the authors and not necessarily those of the NHS, the NIHR or the Department of Health and Social Care. The authors would like to acknowledge the assistance of UKOSS reporting clinicians, the UKOSS Steering Committee and The NIHR Clinical Research Networks without whose support this research would not have been possible.

