## Supplementary Figure 1 and 2 for "Impact of SARS-CoV-2 variant on the severity of maternal infection and perinatal outcomes: Data from the UK Obstetric Surveillance System national cohort"

8330 cases notified

382 (4.6%) Data not yet received

1036 (12.4%) Do not meet case definition

6912 (83.0%) Pregnant women admitted with virology confirmed SARS-CoV-2

3371 (49%) Symptomatic and included

3541 (51%) Asymptomatic and excluded

**Supplementary Figure 1: Women included in study from 1^st^ March 2020 to 11^th^ July 2021**

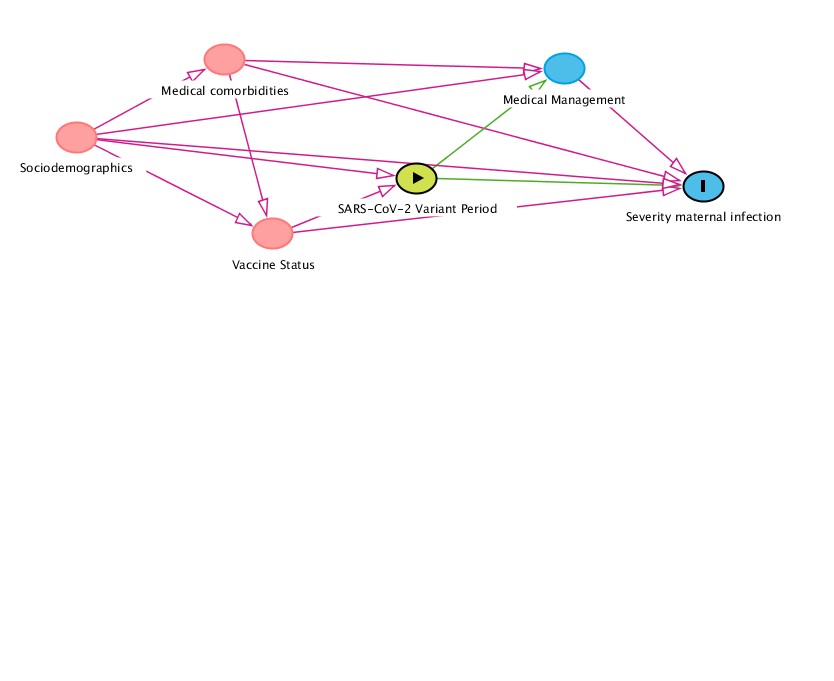

**Supplementary Figure 2: Directed acyclic graphs indicating covariates and mediators on relationship between SARS-CoV-2 variant and severity of maternal infection.**
